# War, Diets, and Mental Health: PTSD in Ukrainian Youth

**DOI:** 10.1101/2025.07.16.25331658

**Authors:** Iryna Halabitska, Pavlo Petakh, Mykhailo Buchynskyi, Iryna Kamyshna, Oleh Lushchak, Oleksandr Kamyshnyi

## Abstract

The ongoing war in Ukraine has exposed young adults to sustained psychological stress, elevating their risk of developing post-traumatic stress disorder (PTSD). In a case–control study of 698 individuals, we investigated associations between PTSD, dietary patterns, disordered eating behaviours, and hematological parameters. PTSD was associated with greater adherence to restrictive diets—including ketogenic, low-fat, and intermittent fasting patterns—as well as higher scores for emotional, external, and uncontrolled eating. Conversely, adherence to a Mediterranean diet was associated with a reduced likelihood of PTSD. Hematologically, PTSD was linked to lower hemoglobin and red blood cell counts, along with elevated inflammatory markers, particularly an increased neutrophil-to-lymphocyte ratio (NLR). Using machine learning, we identified NLR, white blood cell count, and hemoglobin concentration as the strongest predictors of PTSD status. War-related trauma appears to disrupt both eating behaviour and immune function, contributing to the emergence of stress-related psychiatric conditions.

## Main

Since the onset of the full-scale Russian invasion of Ukraine in 2022, young adults residing in the conflict zone have been continuously exposed to high-intensity psychological stressors ^1,2^. This ongoing exposure puts young people at higher risk of posttraumatic stress disorder (PTSD) development, a condition marked by recurring memories of the trauma, avoidance, low mood, and feeling constantly on edge ^3^. While core diagnostic criteria of PTSD focus on affective and cognitive domains, there is increasing recognition that behavioural and somatic dimensions—including lifestyle patterns—may play a critical role in modulating individual responses to trauma ^4^.

Among lifestyle factors, dietary habits and eating behaviours have garnered growing attention due to their potential to both reflect and influence psychological well-being ^5^. Diet is a modifiable behavioural domain that may serve as a coping mechanism under chronic stress, and at the same time exert downstream effects on neuroimmune pathways ^6^. Recent studies suggest that specific diets—like those that cause inflammation or are very restrictive—may increase the risk of mental health problems ^7,8^. In contrast, balanced diets like the Mediterranean diet may help protect against stress ^9,10^. However, it isn’t fully understood how trauma, eating habits, and mental health are connected.

Recently, blood-based signs of inflammation and changes in the immune system have gained attention as possible markers of stress-related mental health conditions ^11,12^. One of these, the neutrophil-to-lymphocyte ratio (NLR), may reflect immune imbalance in disorders like post-traumatic stress disorder (PTSD) ^13^. Despite growing interest, there remains a lack of studies exploring the relationship between this biological marker and behavioral factors like diet and eating habits in shaping stress responses and mental health outcomes ^14,15^. Understanding the interaction between psychological symptoms, behaviors, and biological changes is crucial for identifying individuals at higher risk of stress and for informing early intervention strategies ^16,17^. Young adults experience important mental and brain development, which may increase their sensitivity to the combined impact of trauma and lifestyle factors ^18,19^. Combining psychological, behavioral, and biological approaches can provide a more complete understanding of vulnerability and resilience to stress in the setting of armed conflict ^20,21^.

This research was carried out to examine the associations between PTSD symptoms, dietary patterns, disordered eating behaviors, and hematological parameters in young adults who were residing in Ukraine at the onset of the war, and to assess whether these factors may hold predictive value for stress-related vulnerability.

## Results

### Characteristics of the study participants

The study included 698 participants who were residing in Ukraine at the time of the war’s onset. All participants were categorized into two groups based on the DSM-5 criteria for the presence or absence of PTSD: the Non-PTSD group (n = 250) and the PTSD group (n = 448) (**Fig. 1H**). According to the DSM-5 subscale scores, the Non-PTSD group had a median score of 6 (IQR: 5–9) on the B criterion, compared to 9 (IQR: 7–11) in the PTSD group (p < 0.001; **Fig. 1A**). For the C criterion, the scores were 4 (IQR: 3–5) in both groups (p < 0.001; **Fig. 1B**). On the D criterion, the Non-PTSD group scored 8 (IQR: 6–10), while the PTSD group scored 12 (IQR: 10–14) (p < 0.001; **Fig. 1C**). For the E criterion, the scores were 8 (IQR: 5–10) and 11 (IQR: 9–13.5), respectively (p < 0.001; **Fig. 1D**). The total DSM-5 score was 27 (IQR: 24–29) in the Non-PTSD group and 36 (IQR: 33–39) in the PTSD group (p < 0.001; **Fig. 1E**).

**Figure 1.**
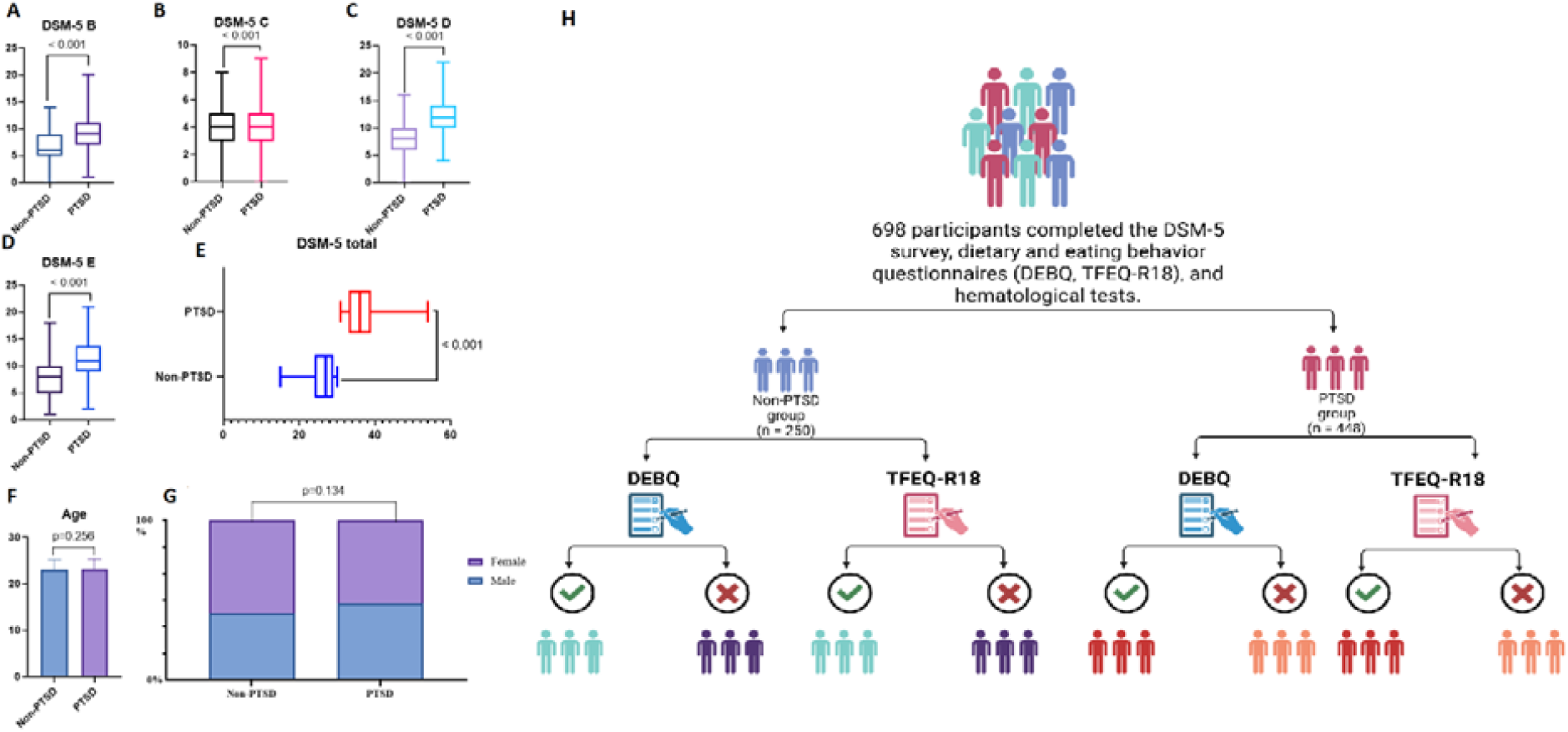
Demographic and symptom characteristics by PTSD status. (A-E) Scores on the DSM-5 PTSD symptom clusters B–E and total scale. (A) Age (years). (G) Gander. The data are presented as median (IQR) and percentages (frequency); DSM-5, Diagnostic and Statistical Manual of Mental Disorders, Fifth Edition; PTSD, posttraumatic stress disorder. (H) Study sample and assessment flow based on PTSD status and completion of eating behavior questionnaires.

The main characteristics of the groups are presented in Figure 1. The age distribution in the Non-PTSD group was 23.0 years (IQR: 21.0–25.0), while in the PTSD group it was 23.0 years (IQR: 22.0–25.0), with no statistically significant difference between the groups (p = 0.256; Fig. 1F). The gender distribution in the Non-PTSD group was 42% male and 58% female, compared to 48.2% male and 51.8% female in the PTSD group, also showing no significant difference (p = 0.134; **Fig. 1G**).

A dietary intake survey revealed a diverse range of eating patterns among participants over the past six months, including no specific diet, a high-protein diet, intermittent fasting, a ketogenic diet, a low-fat diet, a Mediterranean diet, and a vegetarian or vegan diet (**Table 1**). A significantly higher proportion of participants in the PTSD group reported adherence to intermittent fasting (p = 0.037), a ketogenic diet (p = 0.003), and a low-fat diet (p = 0.007). In contrast, a significantly greater number of individuals in the non-PTSD group followed a Mediterranean diet (p < 0.001) (**Table 1**).

**Table 1.** PTSD-Related Differences in Dietary Choices, Eating Behavior Measures, and Hematological Parameters.

| Indicator | Non-PTSD group (n = 250) | PTSD group (n = 448) | p-value |
| --- | --- | --- | --- |
| No specific diet, n(%) | 154 (61.6%) | 218 (48.7%) | 0.074 |
| High-protein diet, n(%) | 9 (3.6%) | 12 (2.7%) | 0.498 |
| Intermittent fasting, n(%) | 14 (5.6%) | 48 (10.7%) | **0.037** |
| Keto, n(%) | 16 (6.4%) | 66 (14.7%) | **0.003** |
| Low-fat diet, n(%) | 5 (2.0%) | 31 (6.9%) | **0.007** |
| Mediterranean diet, n(%) | 34 (13.6%) | 21 (4.7%) | **< 0.001** |
| Vegetarian-vegan, n(%) | 18 (7.2%) | 52 (11.6%) | 0.065 |
| DEBQ-RE, score | 3.1 (2.6-3.5) | 3.1 (2.6-3.6) | 0.07 |
| DEBQ-EE, score | 3.0 (2.5-3.5) | 3.1 (2.5-3.6) | 0.27 |
| DEBQ-ExE, score | 2.7 (2.3-3.2) | 2.9 (2.4-3.4) | **0.003** |
| TFEQ-R18-UE, score | 66.0 (59.0-72.0) | 67.0 (61.0-74.0) | **0.046** |
| TFEQ-R18-EE, score | 65.0 (58.0-71.0) | 67.0 (59.0-73.0) | **0.034** |
| TFEQ-R18-CR, score | 64.0 (57.0-70.0) | 65.0 (59.0-71.0) | 0.251 |
| Hb, g/L | 133.0 (125.0-140.0) | 128.5 (120.0-139.0) | **0.004** |
| RBC, $\times 10^{12}/L$ | 4.3 (4.0-4.7) | 4.1 (3.8-4.6) | **0.016** |
| HCT, % | 40.0 (36.0-42.0) | 38.0 (36.0-41.0) | **0.003** |
| MCV, fL | 85.0 (82.0-88.0) | 84.0 (81.0-88.0) | **0.048** |
| MCH, pg | 28.2 (26.8-28.9) | 27.6 (26.9-28.3) | **0.001** |
| MCHC, g/L | 320.0 (312.0-326.0) | 310.0 (292.0-322.0) | 0.094 |
| Ret, % | 1.0 (0.7-1.1) | 1.0 (0.7-1.2) | 0.627 |
| WBC, $\times 10^9/L$ | 6.3 (6.0-7.0) | 6.65 (5.9-7.2) | **0.045** |
| NEU, $\times 10^9/L$ | 3.4 (3.2-3.9) | 3.6 (3.2-4.1) | **0.006** |
| LYM, $\times 10^9/L$ | 2.2 (2.1-2.4) | 2.2 (1.9-2.3) | **0.002** |
| MONO, $\times 10^9/L$ | 0.42 (0.4-0.5) | 0.45 (0.4-0.5) | 0.224 |
| EOS, $\times 10^9/L$ | 0.1 (0.1-0.2) | 0.1 (0.1-0.3) | 0.058 |
| BASO, $\times 10^9/L$ | 0.07 (0.05-0.1) | 0.08 (0.05-0.1) | 0.093 |
| ESR, mm/h | 10.0 (8.0-13.0) | 11.0 (8.0-15.0) | **0.017** |
| PLT, $\times 10^9/L$ | 234.0 (215.0-248.0) | 230 (205-250) | 0.064 |
| MPV, fL | 9.5 (9.3-10.0) | 9.7 (9.2-10.1) | 0.079 |
| NLR | 1.59 (1.48-1.72) | 1.67 (1.55-1.89) | **< 0.001** |

The data are presented as frequency (percentages) and median (IQR); Differences in diet adherence were assessed by $\chi^2$ test; DEBQ, TFEQ-R18 scores and haematological measures by Mann–Whitney U test. DEBQ, Dutch Eating Behavior Questionnaire; DEBQ-RE, restrained eating; DEBQ-EE, emotional eating; DEBQ-ExE, external eating; TFEQ, Three-Factor Eating Questionnaire; TFEQ-R18, 18-item Revised version of the TFEQ; TFEQ-R18-UE, uncontrolled eating; TFEQ-R18-EE, emotional eating; TFEQ-R18-CR, cognitive restraint; Hb, hemoglobin (g/L); RBC, red blood cells ( $\times 10^{12}/L$ ); HCT, hematocrit (%); MCV, mean corpuscular volume (fL); MCH, mean corpuscular hemoglobin (pg); MCHC, mean corpuscular hemoglobin concentration (g/L); Ret, reticulocytes (%); WBC, white blood cells ( $\times 10^9/L$ ); NEU, neutrophils ( $\times 10^9/L$ ); LYM, lymphocytes ( $\times 10^9/L$ ); MONO, monocytes ( $\times 10^9/L$ ); EOS, eosinophils ( $\times 10^9/L$ ); BASO, basophils ( $\times 10^9/L$ ); ESR, erythrocyte sedimentation rate (mm/h); PLT, platelets ( $\times 10^9/L$ ); MPV, mean platelet volume (fL); NLR, neutrophil-to-lymphocyte ratio.

All participants completed two validated instruments assessing eating behavior: the DEBQ (measuring restrained (DEBQ-RE), emotional (DEBQ-EE), and external eating (DEBQ-ExE)) and the TFEQ-R18 (capturing uncontrolled eating (TFEQ-R18-UE), emotional eating (TFEQ-R18-EE), and cognitive restraint (TFEQ-R18-CR)) (Table 1). Significantly higher scores were observed in the PTSD group for the DEBQ external eating scale (DEBQ-ExE; p = 0.003), the TFEQ-R18 uncontrolled eating scale (TFEQ-R18-UE; p = 0.047), and the TFEQ-R18 emotional eating scale (TFEQ-R18-EE; p = 0.034) **(Table 1)**.

All participants underwent complete blood count analysis (Table 1). The PTSD group exhibited significantly lower levels of hemoglobin (Hb; p = 0.004), red blood cells (RBC; p = 0.016), hematocrit (HCT; p = 0.003), mean corpuscular volume (MCV; p = 0.048), and mean corpuscular hemoglobin (MCH; p = 0.001), alongside significantly higher white blood cell count (WBC; p = 0.045), neutrophils (NEU; p = 0.006), erythrocyte sedimentation rate (ESR; p = 0.017), and neutrophil-to-lymphocyte ratio (NLR; p < 0.001), as well as lower lymphocyte counts (LYM; p = 0.002) **(Fig. 4D)**.

### Dietary patterns, eating behavior disturbances, and hematological parameters influence PTSD development

Participants were stratified into four subgroups according to PTSD status and the presence or absence of disordered eating behaviours, as measured by the Dutch Eating Behavior Questionnaire (DEBQ) and the Three-Factor Eating Questionnaire, 18-item version (TFEQ-R18). Diet type distributions were compared across these subgroups.

In analyses stratified by DEBQ Restraint Eating scores, significant differences in diet type distribution were observed **(Fig. 2A)**. Pairwise comparisons revealed differences between no specific diet and ketogenic diet (p = 0.006), intermittent fasting and Mediterranean diet (p = 0.006), ketogenic and Mediterranean diet (p = 0.002), low-fat and Mediterranean diet (p = 0.002), and Mediterranean and vegetarian/vegan diet (p = 0.019).

**Fig. 2.**
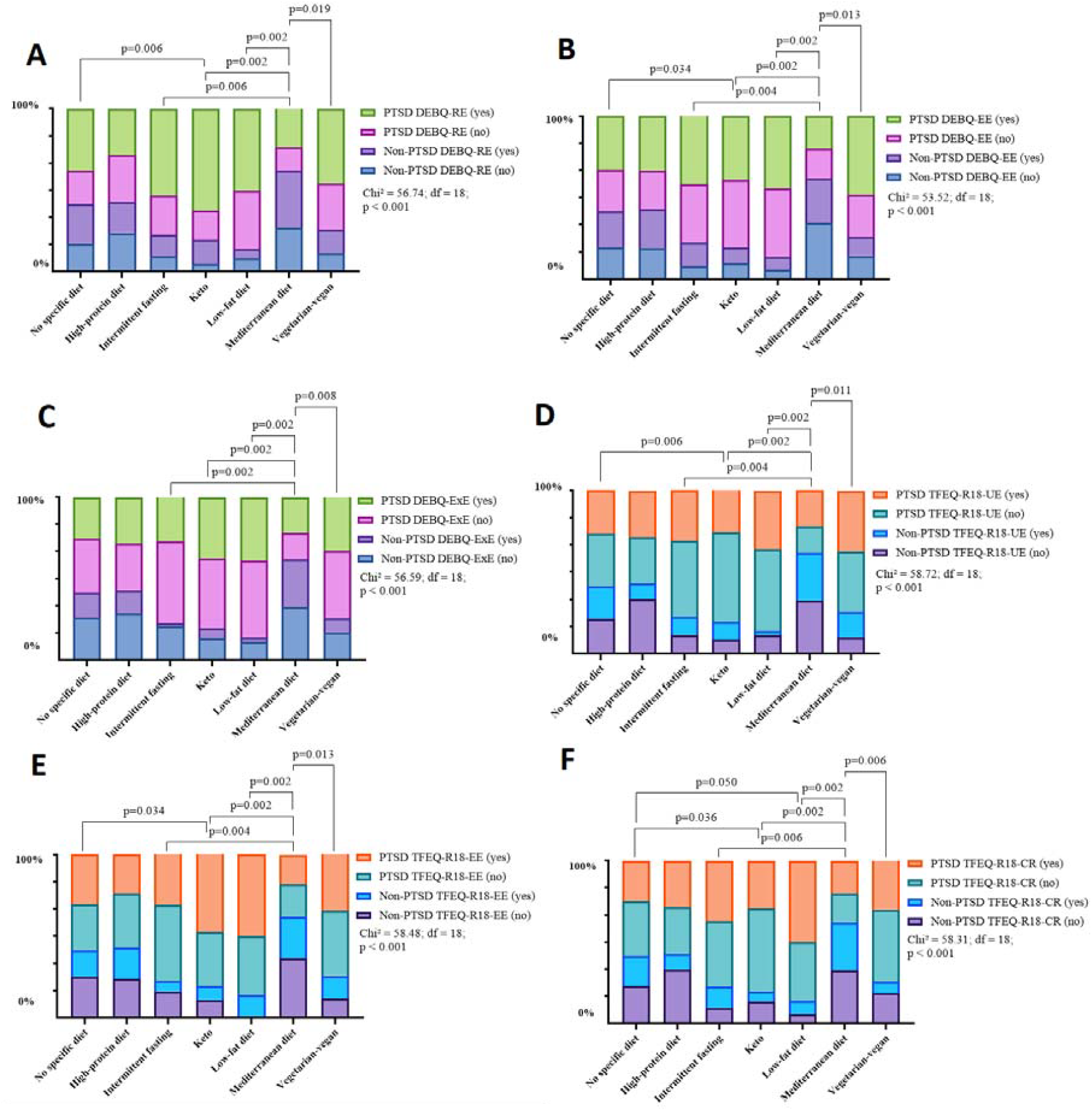
Differences between diet type in PTSD and non-PTSD groups and associated eating behavior scores. (A-F) PTSD and non-PTSD patient groups by DEBQ and TFEQ-R18 score. Data are presented as frequency (percentages); PTSD, posttraumatic stress disorder; DEBQ, Dutch Eating Behavior Questionnaire; DEBQ-RE, restrained eating; DEBQ-EE, emotional eating; DEBQ-ExE, external eating; TFEQ-R18, Three-Factor Eating Questionnaire, 18-item version; TFEQ-R18-UE, uncontrolled eating; TFEQ-R18-EE, emotional eating; TFEQ-R18-CR, cognitive restraint.

A similar pattern was observed in the Emotional Eating subgroup (DEBQ), with significant contrasts between no specific and ketogenic diets (p = 0.034), intermittent fasting and Mediterranean diets (p = 0.004), ketogenic and Mediterranean diets (p = 0.002), low-fat and Mediterranean diets (p = 0.002), and Mediterranean and vegetarian/vegan diets (p = 0.013) **(Fig. 2B)**.

In the External Eating subgroup (DEBQ), differences were again significant, particularly between intermittent fasting and Mediterranean (p = 0.002), ketogenic and Mediterranean (p = 0.002), low-fat and Mediterranean (p = 0.002), and Mediterranean and vegetarian/vegan diets (p = 0.008) **(Fig. 2C)**.

TFEQ-R18-based stratifications yielded comparable findings. Within the Uncontrolled Eating (UE) subgroup, significant differences were found between no specific diet and ketogenic diet (p = 0.006), intermittent fasting and Mediterranean diet (p = 0.004), ketogenic and Mediterranean diet (p = 0.002), low-fat and Mediterranean diet (p = 0.002), and Mediterranean and vegetarian/vegan diet (p = 0.011) **(Fig. 2D)**.

Emotional Eating (EE) scores from the TFEQ-R18 also delineated significant dietary differences, including between no specific and ketogenic diets (p = 0.034), intermittent fasting and Mediterranean diets (p = 0.004), ketogenic and Mediterranean diets (p = 0.002), low-fat and Mediterranean diets (p = 0.002), and Mediterranean and vegetarian/vegan diets (p = 0.013) **(Fig. 2E)**.

Finally, stratification by Cognitive Restraint (CR) revealed significant variation in diet type, with pairwise differences between no specific and ketogenic diets (p = 0.036), no specific and low-fat diets (p = 0.050), intermittent fasting and Mediterranean diets (p = 0.006), ketogenic and Mediterranean diets (p = 0.002), low-fat and Mediterranean diets (p = 0.002), and Mediterranean and vegetarian/vegan diets (p = 0.006) **(Fig. 2F)**.

Dietary patterns varied significantly by PTSD status when examined within strata defined by disordered eating traits, as assessed by the Dutch Eating Behavior Questionnaire (DEBQ) and the Three-Factor Eating Questionnaire (TFEQ-R18).

Restrained eating (DEBQ-RE) was differentially associated with dietary adherence depending on PTSD status. Among individuals without PTSD and low restraint, adherence to no specific diet was significantly lower compared to other groups (p⍰<⍰0.01) and to the ketogenic diet specifically (p⍰<⍰0.01). In contrast, highly restrained non-PTSD individuals more frequently followed Mediterranean (p⍰<⍰0.001) and no specific diets (p⍰<⍰0.05), and less often the ketogenic diet (p⍰<⍰0.05). In those with PTSD, high restraint was associated with lower adherence to no specific (p⍰<⍰0.05) and low-fat diets (p⍰<⍰0.01), and greater use of the ketogenic diet (p⍰<⍰0.001) **(Fig. 3A)**.

**Fig. 3.**
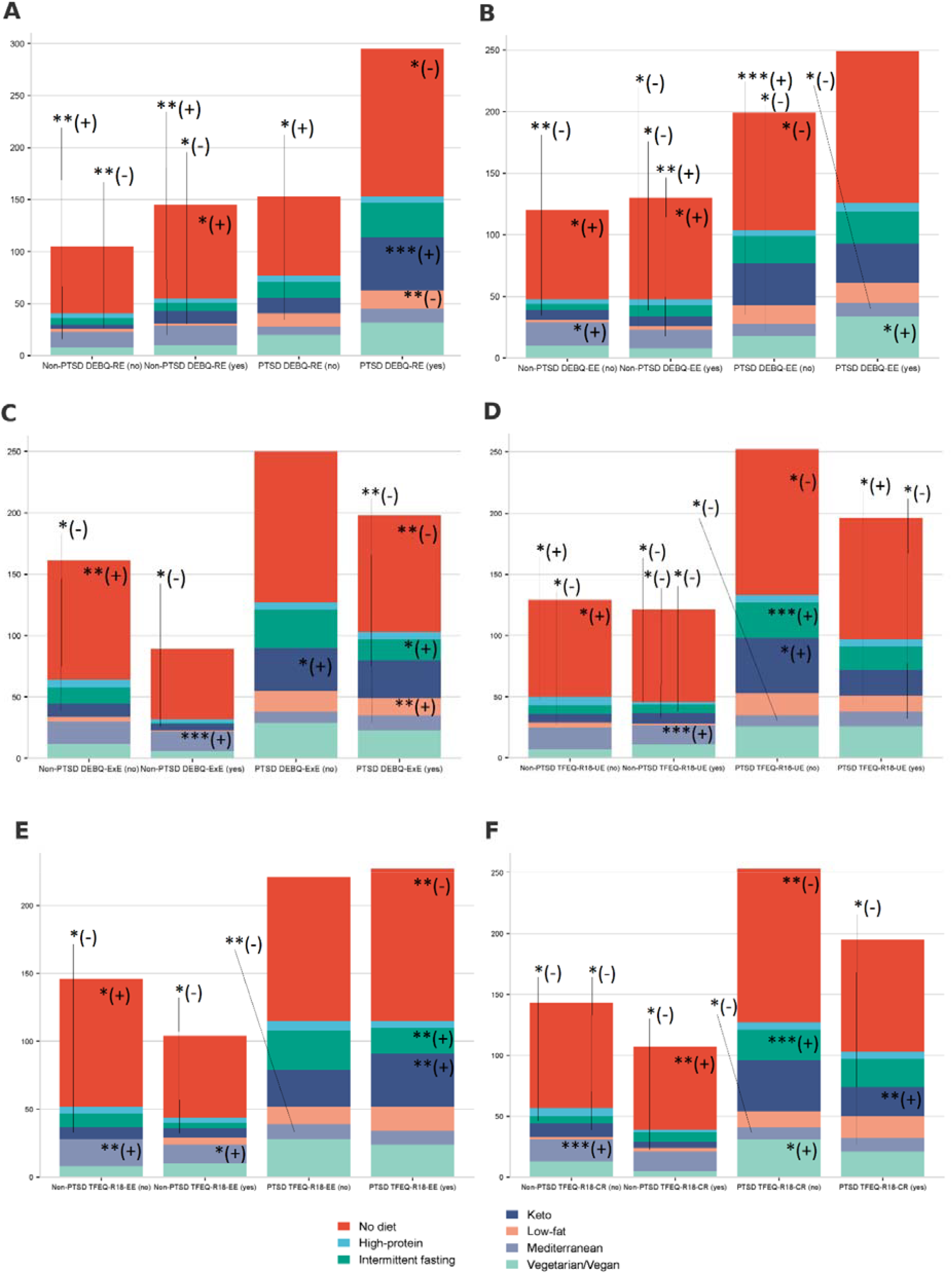
Differences in diet prevalence between PTSD and non-PTSD groups and associated eating behavior scores. (A-F) PTSD and non-PTSD patient groups by DEBQ and TFEQ-R18 score. Data are presented as the number of patients. * p⍰<⍰0.05, ** p⍰<⍰0.01, *** p⍰<⍰0.001. (+) indicates that the diet type is more prevalent in this group compared to others; (−) indicates that the diet type is less prevalent in this group compared to others. PTSD, posttraumatic stress disorder; DEBQ, Dutch Eating Behavior Questionnaire; DEBQ-RE, restrained eating; DEBQ-EE, emotional eating; DEBQ-ExE, external eating; TFEQ, Three-Factor Eating Questionnaire; TFEQ-R18, revised 18-item version; TFEQ-R18-UE, uncontrolled eating; TFEQ-R18-EE, emotional eating; TFEQ-R18-CR, cognitive restraint.

**Fig. 4.**
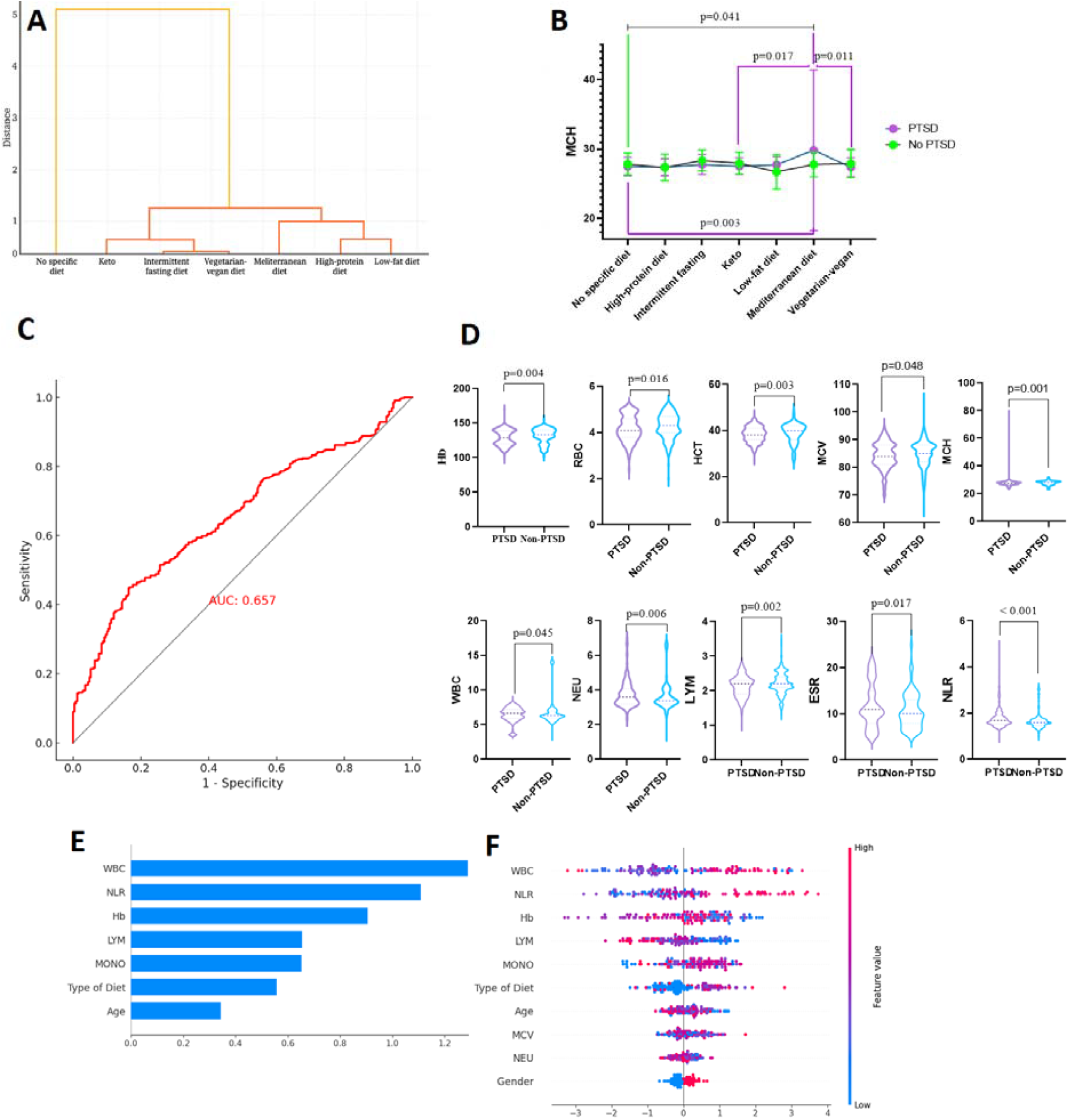
Influence of dietary and hematological factors on PTSD risk and SHAP-based interpretation of XGBoost model predictions. (A) Hierarchical clustering of dietary patterns in relation to PTSD. (B) MCH levels in PTSD and non-PTSD patients across dietary patterns. (C) ROC curve from multivariate model predicting PTSD. (D) Hematological parameters in PTSD and non-PTSD patient groups. (E) Bar plot of mean absolute SHAP values indicating the average impact of each feature on model output; WBC, NLR, and hemoglobin were the most influential predictors. (F) SHAP summary (beeswarm) plot showing distribution and directionality of individual feature effects; higher WBC and NLR were associated with increased PTSD risk. Each point represents one participant, with color indicating the feature value (red = high, blue = low). Higher WBC and NLR values were associated with increased model-predicted PTSD risk.

Emotional eating (DEBQ-EE) also showed PTSD-specific associations. Among non-PTSD participants with low emotional eating, adherence to Mediterranean and no specific diets was higher (both p⍰<⍰0.05), and lower for the ketogenic diet (p⍰<⍰0.01). Those with high emotional eating more often reported Mediterranean (p⍰<⍰0.01) and no specific diet use (p⍰<⍰0.05), and less adherence to ketogenic and intermittent fasting diets (both p⍰<⍰0.05). In PTSD, low emotional eating predicted higher adherence to low-fat diets (p⍰<⍰0.001) and lower use of Mediterranean and no specific diets (both p⍰<⍰0.05), while high emotional eating was associated with vegetarian/vegan diets (p⍰<⍰0.05) **(Fig. 3B)**.

External eating (DEBQ-ExE) further stratified dietary behaviour. Among non-PTSD individuals, both low and high ExE were linked to lower ketogenic diet use (p⍰<⍰0.05), and greater adherence to Mediterranean (p⍰<⍰0.001) or no specific diets (p⍰<⍰0.01). PTSD participants with low ExE showed greater ketogenic adherence (p⍰<⍰0.05), whereas those with high ExE reported increased use of low-fat (p⍰<⍰0.01) and intermittent fasting diets (p⍰<⍰0.05), and reduced adherence to Mediterranean and no specific diets (both p⍰<⍰0.01) **(Fig. 3C)**.

Uncontrolled eating (TFEQ-R18 UE) revealed notable divergences. In non-PTSD individuals, low UE was linked to higher use of Mediterranean and no specific diets (both p⍰<⍰0.05), and less low-fat diet use (p⍰<⍰0.05). High UE was associated with reduced adherence to high-protein, intermittent fasting, and ketogenic diets (all p⍰<⍰0.05), and greater Mediterranean diet adherence (p⍰<⍰0.001). In the PTSD group, low UE corresponded to lower adherence across most diets (p⍰<⍰0.05), whereas high UE predicted reduced use of the Mediterranean and greater reliance on low-fat diets (both p⍰<⍰0.05) **(Fig. 3D)**.

Emotional eating (TFEQ-R18 EE) mirrored the DEBQ-EE findings. Non-PTSD participants with low emotional eating had lower ketogenic (p⍰<⍰0.05), and higher Mediterranean (p⍰<⍰0.01) and no specific diet use (p⍰<⍰0.05). High emotional eating remained associated with lower ketogenic and higher Mediterranean diet adherence (p⍰<⍰0.05). In PTSD, high EE was linked to increased use of intermittent fasting and ketogenic diets, and decreased no specific diet adherence (all p⍰<⍰0.01) **(Fig. 3E)**.

Cognitive restraint (TFEQ-R18 CR) showed opposing trends by PTSD status. Among non-PTSD participants, low CR was associated with lower adherence to intermittent fasting and ketogenic diets (both p⍰<⍰0.05), and greater adherence to the Mediterranean diet (p⍰<⍰0.001). High CR was linked to lower low-fat diet use (p⍰<⍰0.05) and higher no specific diet adherence (p⍰<⍰0.01). In PTSD participants, low CR predicted greater adherence to intermittent fasting and vegetarian/vegan diets (both p⍰<⍰0.05), and lower adherence to Mediterranean (p⍰<⍰0.05) and no specific diets (p⍰<⍰0.01). High CR was associated with increased ketogenic and decreased Mediterranean diet adherence (both p⍰<⍰0.05) **(Fig. 3F)**.

Multivariable logistic regression found a strong link between diet and PTSD. People on ketogenic, low-fat, or intermittent fasting diets had a higher chance of PTSD. This was compared to those without special diets (p < 0.01). Those who followed the Mediterranean diet had a lower risk of PTSD (p < 0.01). This suggests the Mediterranean diet might help protect against PTSD **(Table 2)**.

**Table 2.** Association between dietary patterns and PTSD: Odds Ratios (OR) with 95% confidence intervals from logistic regression analysis (reference: No specific diet). Univariate and multivariate logistic regression analysis of factors associated

| Diet | OR | 95% CI | p-value |
| --- | --- | --- | --- |
| High-protein diet | 0.94 | 0.39–2.29 | 0.895 |

| Intermittent fasting | 2.42 | 1.29–4.55 | 0.0059 |
| --- | --- | --- | --- |
| Keto | 2.91 | 1.63–5.22 | 0.0003 |
| Low-fat diet | 4.38 | 1.67–11.52 | 0.0028 |
| Mediterranean diet | 0.44 | 0.24–0.78 | 0.0052 |
| **Univariate logistic regression analysis** |  |  |  |
| **Variables** | **Model coefficients,**<br>**b ± m** | **p-value** | **Odds ratio (95% CI)** |
| Age | 0.04 ± 0.04 | 0.2612 | 1.04 (0.97–1.12) |
| Gender | 0.25 ± 0.16 | 0.1145 | 1.29 (0.94–1.76) |
| Type of Diet | 0.08 ± 0.04 | 0.0375 | 1.08 (1.00–1.16) |
| WBC | −0.16 ± 0.06 | 0.0046 | 0.85 (0.77–0.95) |
| NEU | 0.15 ± 0.11 | 0.1535 | 1.16 (0.95–1.43) |
| LYM | −0.27 ± 0.13 | 0.0361 | 0.76 (0.59–0.98) |
| MONO | 0.62 ± 0.49 | 0.2036 | 1.86 (0.71–4.85) |
| NLR | 0.89 ± 0.24 | 0.0002 | 2.43 (1.54–3.89) |
| MCV | 0.00 ± 0.01 | 0.8773 | 1.00 (0.98–1.03) |
| Hb | −0.06 ± 0.08 | 0.4762 | 0.94 (0.81–1.09) |
| **Multivariate logistic regression analysis** |  |  |  |
| Type of Diet | 0.08 ± 0.04 | 0.0496 | 1.08 (1.00–1.16) |
| WBC | −0.62 ± 0.12 | <0.0001 | 0.54 (0.42–0.68) |
| LYM | 1.14 ± 0.45 | 0.0116 | 3.13 (1.29–7.61) |
| NLR | 2.81 ± 0.56 | <0.0001 | 16.61 (5.59–49.35) |

Odds ratios (OR) and 95% confidence intervals (CI) are reported for each variable. Univariate and Multivariate analysis includes variables significant at $p \leq 0.05$ in univariate models. PTSD, posttraumatic stress disorder; OR, odds ratio; CI, confidence interval.

Hierarchical cluster analysis grouped dietary patterns based on PTSD rates **(Fig. 4A)**. The “No specific diet” group was common and had a different link to PTSD. Another group included ketogenic, intermittent fasting, and vegetarian/vegan diets. These diets had stricter rules and were linked to higher PTSD rates. A third group had Mediterranean, high-protein, and low-fat diets. This group showed lower or mixed PTSD rates. The results suggest that strict diets are more common in people with PTSD. Balanced diets like the Mediterranean are linked to less PTSD.

Analysis of blood parameters showed significant differences in mean corpuscular hemoglobin (MCH) across diets depending on PTSD status **(Fig. 4B)**. In people without PTSD, MCH differed between those with no specific diet and those on the Mediterranean diet (p = 0.041). In the PTSD group, MCH varied significantly in comparisons between no specific diet vs. Mediterranean (p = 0.003), ketogenic vs. Mediterranean (p = 0.017), and Mediterranean vs. vegetarian/vegan diets (p = 0.011).

Logistic regression analysis identified key predictors of PTSD **(Table 2)**. In univariate models, a higher neutrophil-to-lymphocyte ratio (NLR) was strongly associated with increased odds of PTSD (p = 0.0002), along with lower white blood cell count (WBC; p = 0.0046), and decreased lymphocyte levels (LYM; p = 0.0361). A weaker association was observed for diet type (p = 0.0375).

In the multivariate model, NLR remained the most robust predictor (p < 0.0001), followed by WBC (p < 0.0001), LYM (p = 0.0116), and diet type (p = 0.0496).

The model showed moderate discriminative performance, with an AUC of 0.657 (95% CI: 0.619– 0.696); sensitivity and specificity were 0.449 and 0.836, respectively **(Fig. 4C)**.

The SHAP bar plot demonstrated that white blood cell count (WBC), neutrophil-to-lymphocyte ratio (NLR), and hemoglobin (Hb) were the most influential features for PTSD prediction in the XGBoost model **(Fig. 4E)**.

The SHAP summary plot visualizes the distribution **(Fig. 4F)**. White blood cell count (WBC), neutrophil-to-lymphocyte ratio (NLR), and hemoglobin (Hb) were the top predictors, having the highest average SHAP values (1.29, 1.11, and 0.90), showing they had the strongest impact on the model. Lymphocyte count (LYM), monocyte count (MONO), and diet type also contributed moderately (SHAP values: 0.65, 0.65, and 0.56). Demographic features such as age, gender, and mean corpuscular volume (MCV) had a lower impact. The SHAP summary plot further showed that higher WBC and NLR values were consistently associated with increased predicted risk of PTSD.

## Discussion

This study reveals a multidimensional interplay between PTSD symptomatology, dietary patterns, disordered eating behaviours, and peripheral blood markers in young adults exposed to war-related trauma. The findings suggest that PTSD is not only characterized by affective and cognitive symptoms but also manifests through distinctive behavioural and physiological signatures that may have diagnostic and prognostic implications. Some previous studies have explored these aspects ^22,23^.

Participants with PTSD more frequently adhered to restrictive dietary patterns such as ketogenic, low-fat, and intermittent fasting regimens. These patterns have been previously associated with efforts to exert control under psychological distress ^24,25^ and may reflect attempts to self-regulate internal emotional states. In contrast, the Mediterranean diet—known for its balanced nutrition and anti-inflammatory effects ^26,27^ —was much less common among individuals with PTSD. Logistic regression confirmed that following this diet was linked to a lower risk of PTSD. This supports earlier findings showing that the Mediterranean diet may benefit mental health and reduce inflammation in the body ^28,29^.

Notably, participants with PTSD also exhibited elevated emotional, external, and uncontrolled eating behaviours. Such patterns align with models of dysregulated stress reactivity and impaired reward processing, which are common in PTSD ^4,30^. The stratified analysis revealed that these maladaptive behaviours often co-occur with restrictive diets, suggesting that attempts at dietary control may coexist with vulnerability to emotional cues and loss of eating regulation—a paradox frequently observed in trauma-exposed populations ^30,31^.

PTSD was associated with specific changes in blood markers, including lower hemoglobin and red blood cell values, and higher white blood cell counts, neutrophil counts, and the neutrophil-to-lymphocyte ratio (NLR). These changes may indicate that the immune system is more active and experiencing increased inflammation, a typical response to stress ^32,33^. Of all the blood markers, NLR was the best at showing who had PTSD, making it a possible sign of how the body reacts to stress, this pattern reflects previously reported outcomes ^34,35^. Combining behavior and blood data could help create better tools for early risk detection ^36,37^.

Psychological trauma engages psychological and physiological regulation systems. This interaction is increasingly recognised as a central focus in translational psychiatry ^38^. Specific nutrient patterns may influence several biological systems. They can affect brain-derived neurotrophic factor (BDNF) levels ^39^. They also shape the composition of the gut microbiota. In addition, they may alter systemic cytokine activity ^40^.

Moreover, the convergence of dietary restriction and dysregulated eating behaviours observed in this study is consistent with dual-process models of trauma coping ^41^. These models propose that trauma-exposed individuals may simultaneously engage in rigid control strategies while remaining vulnerable to impulsive or emotionally driven behaviour ^42^. The paradoxical co-occurrence of fasting and emotional overeating in our sample suggests a complex attempt at self-regulation under stress, which sociocultural and gendered expectations around food, control, and body image may further shape. These dynamics may also have implications for treatment adherence and response in nutritional or psychotherapeutic interventions.

Notably, the identification of immune-related blood markers as reliable predictors of PTSD supports a growing recognition of inflammation as a transdiagnostic mechanism across psychiatric conditions ^43^. Elevated NLR and WBC counts are linked to stress reactivity ^44^. They are also associated with long-term cardiometabolic risk ^45^. Dietary behaviour may serve as a modifiable risk factor and as an indirect marker of psychological strain ^46^. At the same time, immune and blood markers give measurable signs that may help support PTSD diagnosis, alongside questionnaires ^47^.

## Conclusion

This study shows that young adults with PTSD more often follow restrictive diets like ketogenic, low-fat, or intermittent fasting, while the Mediterranean diet was linked to lower PTSD risk. Emotional and uncontrolled eating were also more common in the PTSD group. PTSD was associated with lower hemoglobin levels, altered red cell indices, higher white blood cell and neutrophil counts, and an elevated neutrophil-to-leukocyte ratio (NLR). Logistic regression and Machine learning confirmed NLR, WBC, and hemoglobin as key predictors of disease presence. These results suggest that diet, eating behavior, and blood markers may help identify the PTSD risk and guide future prevention strategies.

## Methods

### Ethics Statement

The research protocol was reviewed and approved by the Ethics Committee of I. Horbachevsky Ternopil National Medical University, Ministry of Health of Ukraine (Protocol No. 81, dated April 3, 2025). The study was conducted in full compliance with the ethical principles outlined in the Declaration of Helsinki (1975 revision). All individuals were provided with detailed information regarding the study’s objectives and procedures, and written informed consent was obtained from each participant.

### Study design

This pilot case–control study included 698 young adult participants residing in Ukraine at the onset of the war. All individuals completed standardized psychological assessments, including evaluation of PTSD symptoms according to DSM-5 criteria, with a cutoff score of >30 used to classify participants into PTSD and non-PTSD groups. Participants reported following various diets, including no specific diet, high-protein, intermittent fasting, ketogenic, low-fat, Mediterranean, and vegetarian-vegan. Eating behaviors were assessed using the Dutch Eating Behavior Questionnaire (DEBQ) and the 18-item Revised Three-Factor Eating Questionnaire (TFEQ-R18). Peripheral blood samples and psychological questionnaires were collected concurrently at a single time point.

Inclusion criteria were young adults living in Ukraine when the war began. They had to give informed consent and complete PTSD assessments based on DSM-5. Exclusion criteria included refusal to participate and history of severe psychiatric disorders other than PTSD, such as schizophrenia or bipolar disorder. Severe physical or chronic illnesses affecting blood parameters were also reasons for exclusion. People with incomplete data or those taking medications that could affect immune or blood tests were excluded to reduce confounding factors.

### Psychological, dietary, and blood assessments

PTSD symptoms were evaluated using a standardized questionnaire based on DSM-5 criteria ^48^. The instrument evaluated symptom presence and severity across diagnostic domains: Criterion A (exposure to traumatic event, defined as residency in Ukraine at the war’s onset), Criterion B (intrusion symptoms), Criterion C (avoidance behaviors), Criterion D (negative alterations in cognition and mood), and Criterion E (alterations in arousal and reactivity). Each domain was scored, and a total severity score was calculated. Participants scoring above a cutoff of 30 were classified as PTSD-positive and stratified accordingly.

Dietary patterns were assessed through self-reported adherence to predefined categories, including no specific diet, high-protein, intermittent fasting, ketogenic, low-fat, Mediterranean, and vegetarian-vegan diets for the last 6 months. Eating behaviors were further characterized using validated questionnaires: the Dutch Eating Behavior Questionnaire (DEBQ) ^49^, assessing restrained eating (DEBQ-RE), emotional eating (DEBQ-EE), and external eating (DEBQ-ExE); and the 18-item Revised Three-Factor Eating Questionnaire (TFEQ-R18), evaluating uncontrolled eating (TFEQ-R18-UE) ^50^, emotional eating (TFEQ-R18-EE), and cognitive restraint (TFEQ-R18-CR). Subscale scores were calculated according to standardized protocols, with scores above the 75th percentile indicating elevated or problematic eating behaviors.

Peripheral blood samples were obtained via venipuncture and analyzed using the Sysmex XN-1000 automated hematology analyzer (Sysmex Corporation, Kobe, Japan) to quantify hemoglobin, RBC, hematocrit, MCV, MCH, MCHC, reticulocytes, WBC, neutrophils, lymphocytes, monocytes, eosinophils, basophils, platelets, and mean platelet volume. Neutrophil-to-lymphocyte ratio was calculated from absolute counts. Erythrocyte sedimentation rate was measured using the Westergren method with an ESR analyzer (Alifax, Padova, Italy). All procedures followed manufacturer protocols and quality controls.

### Statistical analysis

Statistical analyses were performed using IBM SPSS Statistics (version 28.0), GraphPad Prism (version 10), R (version 4.3.1; R Foundation for Statistical Computing), and Python (version 3.10) with the libraries pandas, numpy, scikit-learn, xgboost, and shap. Data visualizations were generated using GraphPad Prism, ggplot2 (R), and seaborn/matplotlib (Python). The Shapiro–Wilk test was used to assess normality of continuous variables. Given non-normal distributions, continuous data are reported as median and interquartile range (IQR), and group comparisons (PTSD vs non-PTSD) were performed using the two-tailed Mann–Whitney U test. Categorical variables, including dietary adherence patterns, were compared using the Chi-square (χ^2^) test of independence. Standardized residuals were examined post hoc to identify cell-level contributions, with absolute values ≥ 1.96, ≥ 2.58, and ≥ 3.29 corresponding to significance thresholds of P⍰<⍰0.05, P⍰<⍰0.01, and P⍰<⍰0.001, respectively. To assess the effect of dietary patterns on hematological parameters within PTSD and non-PTSD groups, we applied the Kruskal–Wallis test followed by Dunn’s multiple comparisons test with Bonferroni correction. Hierarchical cluster analysis was performed to group dietary patterns based on eating behavior questionnaire responses, using Ward’s linkage and squared Euclidean distance. Resulting clusters were evaluated for associations with PTSD status. Univariate and multivariate logistic regression were used to identify independent predictors of PTSD. Model performance was evaluated using ROC curves and AUC values. A supervised machine learning approach was implemented using an XGBoost classifier. Continuous variables were normalized via quantile transformation. The dataset was split into training (80%) and test (20%) subsets. SHAP values (TreeExplainer) were used to explain the model. Feature importance was shown with bar and beeswarm plots.

## Data Availability

All data produced in the present study are available upon reasonable request to the authors

## Data availability

Data will be made available on request to the corresponding author/s.

## Code availability

Codes for software used in this analysis are all publicly available through the citations for each method, as introduced.

## Acknowledgements

none to declare.

## Author contributions

Conceptualization, O.K.; collected and analyzed the data, I.H., P.P., M.B., O.K.; writing—original draft preparation, I.H., P.P., M.B., O.K.; writing—review and editing, I.H., P.P., M.B., I.K., O.L., O.K.; visualization, I.H., P.P., M.B., O.K.; supervision, O.K. All authors have read and agreed to the published version of the manuscript.

